# Dysfunctional HDL Promotes Platelet Apoptosis and Thrombosis in Familial Hypercholesterolemia

**DOI:** 10.1101/2025.03.26.25324730

**Authors:** Karthik Dhanabalan, Honglei Li, Patricia G. Yancey, Sergey Solomevich, Jiyu Li, Yanshuang Li, Jiansheng Huang, Connor Dennewitz, Chunyan Wang, Huan Tao, Loren Smith, David Gailani, Daria Salamevich, Kejun Shao, Jing Du, Kathleen Martin, John Hwa, Sean S. Davies, MacRae F. Linton, Wen-Liang Song

## Abstract

**Background:** In familial hypercholesterolemia (FH), high-density lipoprotein (HDL) often becomes dysfunctional and enriched with lipid peroxidation products, potentially contributing to increased thrombotic risk. However, its specific effects on platelet function and thrombosis remain unclear. Whether targeting HDL oxidation can restore its protective role has yet to be determined.

**Methods:** Platelet function in healthy and FH subjects was assessed via flow cytometry, TEM, western blotting, and transcriptome analysis. The effects of HDL from healthy and FH subjects and lipid peroxidation-modified HDL on oxidized low-density lipoprotein (oxLDL)-induced platelet activation and apoptosis were evaluated. *In vivo* thrombosis was assessed in LDL-receptor-deficient (*Ldlr*^-/-^) mice fed a Western-style diet and treated with 2-hydroxybenzylamine (2-HOBA), a lipid peroxidation scavenger. The roles of SR-B1 and CD36 in platelet activation were examined using inhibitors.

**Results:** Platelet activity was elevated in FH subjects compared to healthy controls, with FH platelets showing increased apoptosis, higher pro-apoptotic and reduced anti-apoptotic proteins. HDL from healthy subjects attenuated oxLDL-induced platelet activation and apoptosis, whereas FH-HDL exacerbated these effects. Western blot and immunofluorescence confirmed that control HDL prevented platelet activation, while FH-HDL promoted apoptosis in oxLDL-stimulated platelets. FH-HDL was enriched with peroxidation products, and lipid peroxidation-modified HDL from healthy volunteers exhibited similar pro-apoptotic effects. Treatment with 2-HOBA mitigated dysfunctional HDL-induced apoptosis, improved thrombosis outcomes, and enhanced blood flow in *Ldlr*^-/-^ mice. Blocking SR-B1 abolished the protective effects of healthy HDL but had no impact on FH-HDL, whereas inhibiting CD36 prevented the pro-apoptotic effects of FH-HDL.

**Conclusion:** Our research shows that while HDL normally protects against platelet apoptosis, in familial hypercholesterolemia it turns prothrombotic, enhancing platelet dysfunction and thrombosis. Treatment with 2-HOBA effectively counters these adverse effects, highlighting a potential therapeutic strategy for managing cardiovascular risks in FH patients.

## Introduction

Familial hypercholesterolemia (FH) is a dominantly inherited genetic disorder resulting from mutations in genes associated with low-density lipoprotein (LDL) receptor function, leading to substantially raised levels of LDL cholesterol and a heightened susceptibility of early-onset atherosclerosis and cardiovascular events^1,2^. Despite advances in LDL-lowering therapies, individuals with FH continue to experience residual cardiovascular risk, suggesting that factors beyond LDL cholesterol levels contribute to disease progression^3^. Dysfunctional high-density lipoprotein (HDL) has emerged as a critical area of investigation, potentially influencing cardiovascular outcomes in FH patients.

HDL is traditionally known for its atheroprotective properties, including its function in reverse cholesterol transport, antiapoptotic activity and antithrombotic effects^4^. However, emerging evidence suggests that HDL can undergo structural modifications in response to oxidative stress and metabolic dysfunction, leading to impaired functionality. One such modification is the interaction of Apolipoprotein (apo) A1, a major protein component of HDL with dicarbonyl compounds, including malondialdehyde (MDA). The resulting apoA1 dicarbonyl modification alters its tertiary structure and diminishes its capacity to effectively mediate cholesterol efflux, prevent oxidation, and promote cell survival, thereby contributing to platelet activation, plaque instability, and increased susceptibility to thrombosis^5^.

Mitochondria are well-documented in nucleated cells, where their functions are well understood. However, their significance amplifies within the nucleus-free environment of platelets. Mitochondria are crucial regulators of cell fate, contributing not only to energy production but also to apoptotic pathways by releasing factors like cytochrome C^6^. In human blood platelets, mitochondria serve as efficient energy producers for both resting and activated states. They also participate in non-ATP-dependent functions, including generating reactive oxygen species and influencing apoptotic-like processes. Damage to mitochondria is pivotal in platelet apoptosis, leading to the loss of mitochondrial membrane potential (ΔΨm) under various conditions including storage^7^. Alterations in the mitochondrial permeability transition pore are proposed to contribute to the formation of coated platelets-a subset observed after agonist activation (thrombin or collagen) which express phosphatidylserine (PS) on their surface. Therefore, mitochondrial integrity and function play crucial roles in regulating platelet activation and apoptosis, processes that share common features such as PS externalization^8^.

Platelet activation and apoptosis are fundamental processes governing the intricate balance between hemostasis and thrombosis. Platelets possess intricate mechanisms for lipid uptake and metabolism^9^. Scavenger receptor class B type 1 (SR-B1) and cluster of differentiation 36 (CD36) receptors on platelets enable the recognition and uptake of lipoproteins, including HDL. These receptors not only facilitate lipid transport but also modulate platelet activation and function^10^. SR-B1, originally identified for its function in cholesterol metabolism, has been shown to alter platelet activation by binding to lipoproteins and facilitating lipid transport^11^. CD36, known for its function in lipid absorption and inflammation, serves as a receptor for various ligands, including oxidized low-density lipoproteins and thrombospondin-1, contributing to platelet activation and adhesion at sites of vascular injury^12^. Dysfunctional HDL, characterized by alterations in composition and function, can disrupt lipid metabolism via SR-B1 and CD36 receptors, potentially contributing to thrombotic events. Therefore, we conducted an investigation to determine whether dysfunctional FH-HDL could induce mitochondrial dysfunction and impact platelet destiny. Our findings revealed that the dysfunctional FH-HDL-mediated impact on mitochondrial function plays a dual role by influencing both platelet activation and apoptosis, effects that are due to altered interaction with SR-BI and CD36.

## Methods

### Mice

*Ldlr^−/−^* and WT mice on a C57BL/6 background were sourced from the Jackson Laboratory. All animal studies adhered to the relevant ethical guidelines for vertebrate animal research. The protocols for the animals were approved by Vanderbilt University’s Institutional Animal Care and Use Committee and conducted in accordance with its regulations. Mice were housed on either a standard chow diet or a Western-type diet, which consisted of 17 kcal% protein, 43 kcal% carbohydrate, and 40 kcal% fat (Research Diets Inc., NJ, USA). Eight-week-old male *Ldlr^−/−^* mice on a chow diet were pretreated with either vehicle alone (water) or water containing 1 g/L of 2-HOBA (as the acetate salt, Metabolic Technologies, Inc., Ames, IA). After 2 weeks of pretreatment, the mice continued with these treatments while transitioning to a Western diet for 16 weeks to promote hypercholesterolemia and atherosclerosis. Based on the average weight and daily water intake of each mouse, the estimated daily dosage of 2-HOBA at 1 g/L is 200 mg/kg.

### Assessment of Platelet Aggregation

Sodium citrate (3.2%) tubes were used to collect fresh blood from healthy and FH subjects. Platelet-rich plasma (PRP) isolation involved spinning the blood samples at 200 x g and at 1000 x g without braking. Different concentrations of collagen were added to 500µL of blood or PRP and platelet aggregation was measured using a Chronolog model 700 (Chronolog Corp., PA, USA)

### P-selectin levels

P-selectin levels were measured in both whole blood and PRP by ELSIA kit (Invitrogen, CA, USA).

### Human Platelet Preparation

The PRB was centrifuged for washed platelets at 1000 g (25°C, 10 min). Platelets were reconstituted in Tyrode’s buffer consisting of HEPES (5mmol/L), NaCl (137 mmol/L), KCl (2mmol/L), MgCl2 (1 mmol/L), NaHCO3 (12 mmol/L), NaH2PO4 (0.3 mmol/L), glucose (5.5 mmol/L) and BSA (3.5mg/mL). Z1 particle counter (Beckman Coulter, CA, USA) was utilized to determine the platelet number.

### Flow Cytometry Analysis of Platelet Activation, Apoptosis, and ΔΨm

To measure platelet activation, the platelets were treated with anti-CD62P and anti-CD41 (Thermofisher, MA, USA) for 30 minutes at room temperature followed by fixation with 4% paraformaldehyde. The fluorescent signal was analyzed using flow cytometry. For determination of apoptosis and ΔΨm, platelet suspensions were incubated with 40 nM TMRM (Thermofisher, MA, USA), followed by staining with 4 µg/ml annexin V (Thermofisher, MA, USA). Based on their forward and side scatter characteristics, platelets were identified and gated for analysis.

### Transmission Electron Microscopy

Platelets were fixed in 2.5% glutaraldehyde in 0.1M cacodylate buffer, post fixed in OsO4 (1%), and en-block stained with uranyl acetate (1%). The samples were dehydrated using a stepwise approach with varying concentrations of ethanol and then infiltrated with Quetol 651 Spurr’s resin, employing propylene oxide as the transition solvent. The process concluded with polymerization at 60°C for 48 hrs. Specimen sections were cut at a nominal thickness of 70 nm on a Leica UC7 and placed onto 300 mesh Ni grids. Imaging was performed on a Tecnai T12 transmission electron microscope at 100 keV, using an AMT Nanosprint5 CMOS camera, on the samples. The image was split into sections, platelet changes of budding and vacuolization were counted and normalized per total platelet number.

### RNA Sequencing and Gene Expression Analysis

The platelet was reconstituted in 700 µl of QIAzol Lysis Buffer (Qiagen, Hilden, Germany) and stored immediately at-80°C. To avoid batch differences, RNA was extracted from all platelet samples together using the RNeasy Mini Kit (Qiagen, Germany; Cat: 1071023).

NEBNext rRNA Depletion Kit (NEB, Cat: E6310X) were used to prepare RNASeq libraries. Sequencing of the libraries was performed on the NovaSeq 6000 platform, utilizing 150 bp paired end reads with an aim of achieving 50 million reads for all samples. RTA (version 2.4.11; Illumina) was employed for base calling, while the analysis was conducted utilizing MultiQC v1.7. Additionally, the analysis was performed with 3.6.3 version of Dragen Software was employed to align the data with the hg38 or mm10 reference genomes.

Paired-end RNA sequencing reads, each 150 base pairs in length, underwent quality trimming and filtering through Trimgalore v0.6.7^13^. The processed reads were subsequently aligned and quantified using Spliced Transcripts Alignment to a Reference (STAR)^14^ v2.7.9a, employing the –quantMode GeneCounts parameter against the Hg38 human genome. Read counts from the samples were normalized, and differential expression analysis was conducted utilizing DESeq2 v1.36.0^15^. Features that were counted fewer than five times across a minimum of three samples were excluded from further analysis. To visualize the principal component analysis, batch effect regression was applied using Limma v3.54.0^16^. Gene Set Enrichment Analysis (GSEA) was carried out with the R package Clusterprofiler^17^, utilizing gene sets sourced from the Human MSigDB database v2022.1.Hs^18^.

### Western Blotting

End of treatment, protein samples were extracted from platelets, and BCA protein assay kit (Thermo Fisher, MA, USA) was used to estimate the platelet protein concentration. 30ug of protein samples were resolved by SDS-PAGE Electrophoresis. Primary antibodies targeting the proteins listed below were used in this study: T-AKT, pAKT (Thr 308), PI3K p85, Bcl-xL, Bcl-2, Bax, active caspase-3, PKCδ and cytochrome C (Cell Signaling). The blots were reprobed with a β- actin antibody (Cell Signaling) for normalization. Detection and quantification were performed using Image Studio digits software.

### Confocal Microscopy

Following treatment, the washed platelets were permitted to settle on glass-bottom dishes for a duration of 30 minutes. Fixation was performed for 15 minutes using 4% paraformaldehyde in PBS. Subsequent to fixation, the platelets underwent permeabilization for 10 minutes with 3% BSA and 0.5% Triton X-100 in PBS. They were then incubated overnight at 4°C with various primary antibodies, including CD41 (APC conjugated), activated Caspase 3, PKCδ, Cytochrome C, PI3K-p85, and Bcl-xL. Following washing, the platelets were subsequently incubated with Alexa Fluor 488 conjugated anti-rabbit antibodies (Thermofisher, MA, USA). The staining of the platelets was examined utilizing a Zeiss LSM 880 confocal microscope, which was fitted with a 60X oil immersion lens.

### Carotid Artery Occlusion

At the end of the experimental period, mice were anesthetized with isoflurane, and surgical procedures were performed after confirming the depth of anesthesia by toe pinch or eye blink reflex. A midline cervical incision was performed, followed by the placement of a Doppler flow probe (Model 0.5 VB, Transonic Systems, Ithaca, NY) on the right carotid artery. This probe was subsequently linked to a flowmeter (Transonic Model T105), and the resulting data were analyzed using a computerized data collection program, Powerlab (AD Instruments, CO). 0.12 ml of Rose Bengal (Fisher Scientific, Fair Lawn, NJ) injected into the jugular vein at a concentration of 50 mg/kg. Following the injection, the jugular vein was completely ligated to ensure hemostasis. A 1.5-mW green light laser (540 nm) (Melles Griot, Carlsbad, CA) was directed at the carotid artery from 5 cm away for up to 120 minutes or until thrombosis was observed, prior to the injection. Blood flow in the vessel was continuously observed from the initiation of the injury until the conclusion of the experiment. Once total occlusion was achieved, the mice were euthanized with an overdose of isoflurane, then cervical dislocation was performed. The carotid arterial segments that had undergone injury were excised and embedded in paraffin to verify the presence of occlusive thrombosis.

### Tail Bleeding Time

Tail bleeding time was measured using a modified version of an established method^19^. Mice were anesthetized as described earlier and placed on a heating pad set to 37°C. The tail was then transected with a sterile scalpel at a point where the tail’s diameter was approximately 1 mm (2–4 mm from the tip). Immediately following transection, the tail was submerged in a 50-mL Falcon tube containing 0.9% NaCl, pre-warmed to 37°C. The time taken for bleeding to stop was recorded. No mouse was allowed to bleed for more than 30 minutes. The total bleeding time was documented, including cases where rebleeding occurred within the 30-minute window. Mice were euthanized by cervical dislocation immediately after the experiment, before recovering from anesthesia.

### Statistical Analysis

GraphPad Prism 9 software was used to perform the statistical analysis (GraphPad Software, Inc.). All data are expressed as mean ± standard error. Group comparisons were executed using an unpaired Student’s t-test for two groups. A p-value of ≤0.05 was considered statistically significant.

## Results

### Increased platelet activity and apoptosis were observed in FH patients

Similar to previous reports with platelets from hypercholesterolemic subjects^20^, platelets from FH versus control subjects were more reactive to ADP, an aggregating agent, when analyzed using either whole blood or PRP preparations. (Figure 1A). P-selectin (CD62) is a cell adhesion molecule located on the surface of activated platelets, and its levels also were elevated upon addition of ADP in FH subjects’ whole blood or PRP compared to healthy subjects (Figure 1B). Interestingly, TEM analysis revealed that the FH platelets are hyperactive as FH versus control platelets had markedly increased microparticle size, platelet aggregation and pseudopodia elongation (Figure 1C). Thromboxane A2 (TxA2) is a lipid released by platelets and other cells, and its levels in urine can indicate platelet hyperactivation^21^. In this study, we observed that urinary excretion of TxA2 was significantly higher in individuals with FH compared to healthy subjects (Figure 1D). In addition, to assess the platelet apoptosis in FH subjects in comparison to healthy subjects, we evaluated the platelet ΔΨm using TMRM staining and externalization of phosphatidylserine (PS) by annexin V staining using flow cytometry. TMRM is easily taken up by mitochondria and its fluorescence is rapidly lost when ΔΨm is decreased. In FH subjects, the platelet ΔΨm is significantly less than control platelet ΔΨm, which is indicated by TMRM intensity (90±2.8 vs 78.3±1.6). At the same time annexin V levels were elevated (7.3±1.4 vs 18±1.6) in platelets from FH subjects compared to healthy subjects (Figures 1E and 1F). The concurrent rise in platelet aggregation, P-selectin levels, and annexin V binding indicates that a significant portion of the circulating platelet population in patients with FH is experiencing activation and apoptosis.

**Figure 1.**
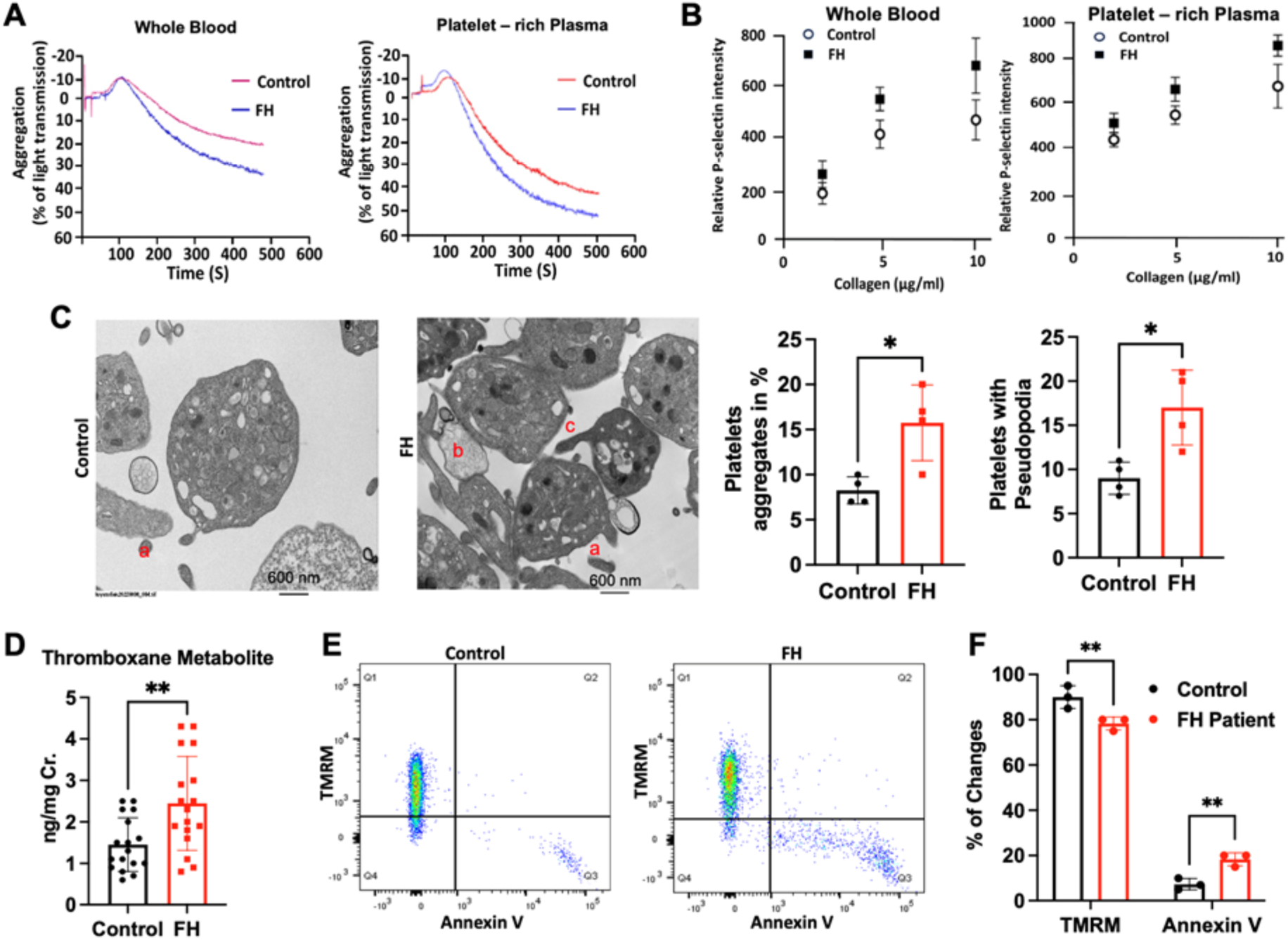
Platelet activity and apoptosis in FH patients reflect increased thrombotic potential. **A,** upon addition of ADP (50µM) the aggregation rate was elevated in both whole blood and PRP from FH subjects compared to healthy individuals. **B,** P-selectin levels also were elevated upon addition of ADP in FH subject’s whole blood or PRP compared to healthy subjects. **C,** Transmission electron microscopy shows that FH patients’ platelets had more aggregation, activation, and budding formation than healthy subjects. **D,** 40nM of TMRM was used to measure mitochondrial membrane potential (ΔΨm) and Annexin V was used to detect phosphatidylserine externalization. **E** and **F,** the bar diagram indicates that in FH subjects, the platelet ΔΨm is less than controls at the same time annexin V levels were elevated in FH compared to healthy individuals. FH: Familial hypercholesterolemia. Data are expressed as mean ± SEM. *- P ≤ 0.05; **- P ≤ 0.01; *** - P ≤ 0.001.

To further characterize platelet activity in FH (N=13) versus control (N=10) subjects, we performed RNAseq (RNA sequencing). The heatmap displays the top 100 genes, ranked by adjusted p value (Figure 2A). In FH subjects, all of the top 100 genes were upregulated compared to the healthy control platelet transcriptome (Figure 2A). Interestingly, the dot plot analysis shows that critical genes involved in apoptosis, coagulation, and inflammation were significantly upregulated in FH patients’ platelets (Figure 2B).

**Figure 2.**
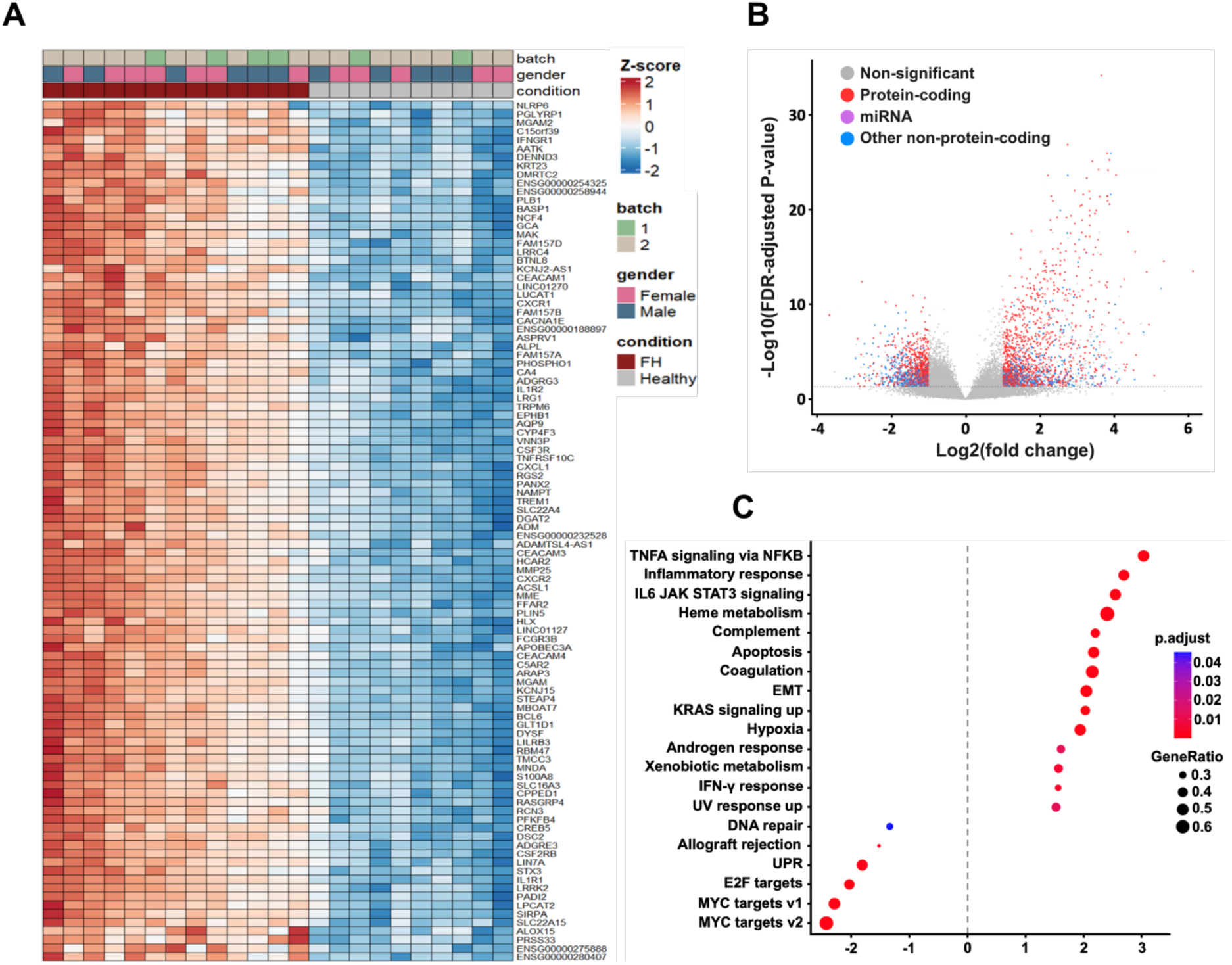
RNA sequencing reveals differential expression of lncRNAs and protein-coding genes in platelets from FH patients. A, Hierarchical clustering map of lncRNAs between FH patients and control samples. **B,** A volcano plot illustrates the relationship between log2 fold change (log2FC) and false discovery rate (FDR)-adjusted P-values, contrasting FH samples with control samples. This visualization highlights differentially expressed protein-coding genes (PCGs), with upregulated genes marked in red and downregulated genes in blue. Additionally, specific PCGs of interest are emphasized for clarity. **C,** Volcano plot indicates the inflammatory response, coagulation and apoptosis pathways are significantly upregulated in FH patients compared with control samples.

### Evidence of mitochondrial damage causes platelet apoptosis

The intrinsic mitochondria-dependent pathway (IMDP) plays a crucial role in the process of programmed cell death, serving as a primary mechanism through which cells can initiate apoptosis. This pathway is controlled by a complex balance between pro-apoptotic and anti-apoptotic proteins. Therefore, we sought to determine the pro-and anti-apoptotic marker levels in platelets obtained from both healthy and FH patients. PKCδ conveys the proapoptotic signal through mitochondrial proteins BAX/BAK, triggering the release of cytochrome c and activating caspase 3. At the same time, PKC stabilizes BCL-xL to suppress the antiapoptotic process^22^. In the present study, we found that the proapoptotic markers PKCδ, BAK, BAX, cytochrome C and activated caspase 3 levels were elevated in platelets-isolated from FH subjects compared to controls by western blotting. In addition, the antiapoptotic proteins PI3K p85, pAKT (Thr 308) and BCL-xL levels were significantly reduced in FH versus control platelets (Figure 3A). Immunofluorescence studies also showed that the expression of the proapoptotic markers, PKCδ, cytochrome C and activated caspase 3 was significantly higher, whereas the antiapoptotic proteins, PI3K p85 and BCL xL were diminished in FH compared to healthy platelets (Figures 3B and 3C). Together, these findings are consistent with enhanced permeabilization of the outer mitochondrial membrane in FH platelets leading to activation of caspase 3 and inhibition of anti-apoptotic proteins thereby promoting apoptosis and platelet activation.

**Figure 3.**
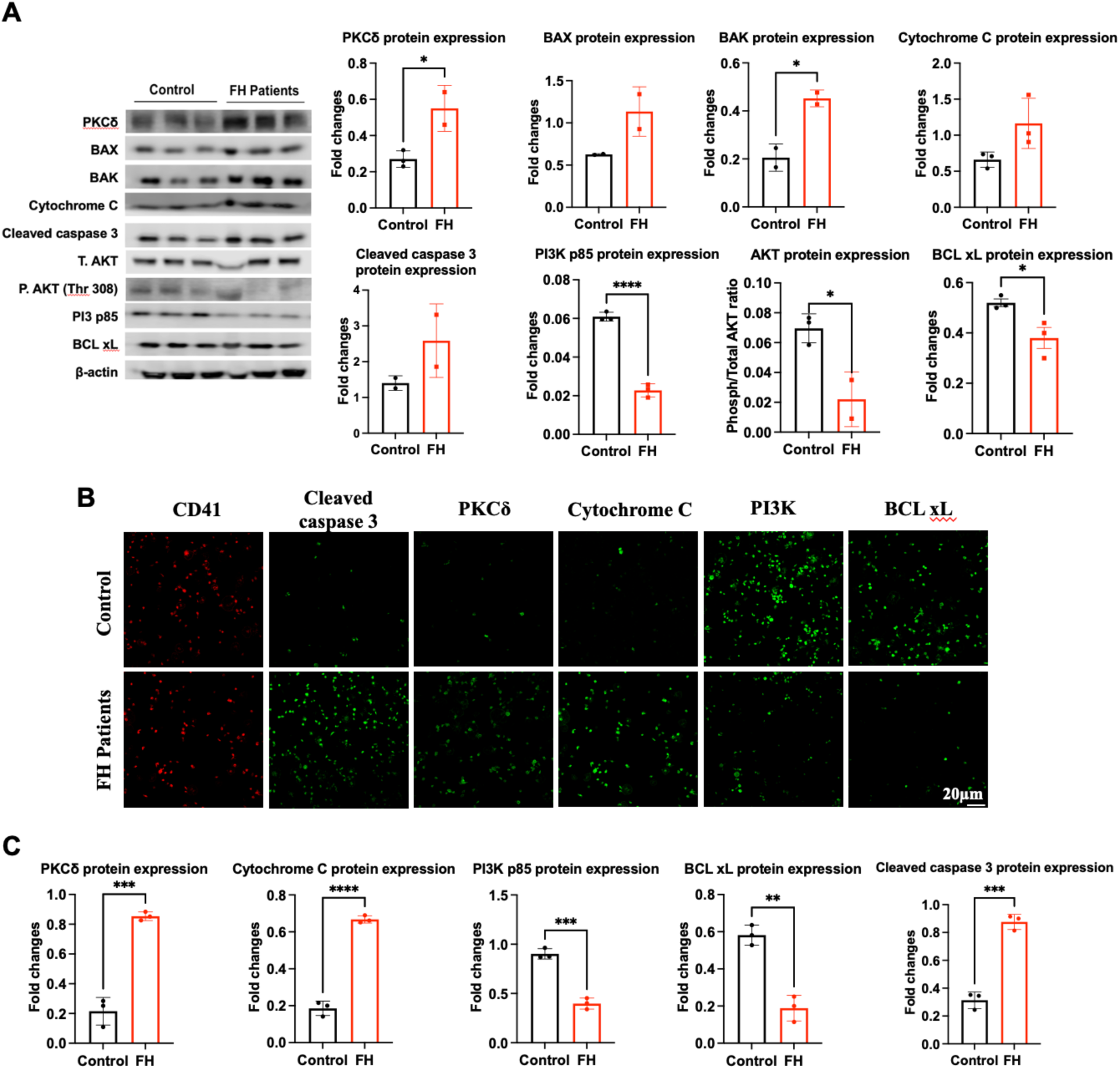
Mitochondrial damage and enhanced apoptosis in platelets from FH patients. A, Representative blots shows the protein expression for PKC delta, BAX, BAK, cytochrome C, activated caspase 3, PI3K p88, pAKT (Thr 308), total AKT, and BCL xL in platelets isolated from healthy and FH subjects. Bar diagram for western blot analysis suggest that apoptotic signaling proteins are elevated in FH patient platelets and anti-apoptotic markers are downregulated in FH patient platelet compared to healthy. **B** and **C,** Representative images from immunostaining and the level of apoptotic markers in platelets isolated from control and FH patients was measured by immunostaining. Data are expressed as mean ± SEM. * - P ≤ 0.05; ** - P ≤ 0.01; *** - P ≤ 0.001.

### FH-HDL is impaired at preventing OxLDL-induced platelet activation and apoptosis

To analyze the role of HDL in FH platelet apoptosis, we isolated the HDL from healthy and FH subjects and administered it to washed platelet preparations treated with OxLDL (oxidized LDL). We used OxLDL as a stimulus because plasma from FH subjects is markedly enriched with OxLDL^23^. To characterize the effect of OxLDL on platelets, apoptosis was examined with increasing doses of OxLDL (50, 100, 250µg/mL). As 100µg/mL of OxLDL induces a moderate amount of apoptosis (12.5±0.25 vs 0.6±0.1) but not a severe degree of death like 250µg/mL (28.5±0.3.5 vs 0.6±0.1) when compared to untreated platelets, the action of HDL on OxLDL induced apoptosis were examined using 100µg/mL of OxLDL (Figure 4A). HDL from control subjects significantly increased the ΔΨm of the platelet population and decreased the number of Annexin-V-positive cells in OxLDL treated platelets. In contrast, HDL from FH subjects did not protect platelets from the OxLDL induced decrease in ΔΨm. In addition, FH-HDL promoted more apoptosis as determined by Annexin-V staining than OxLDL treatment alone, suggesting that FH-HDL accelerates the apoptosis process in platelets treated with OxLDL (Figure 4B). To gain a deeper understanding of how FH-HDL influences platelet apoptosis, we determined the platelet’s morphological features by TEM (Figure 4C). Platelet microparticle size, degranulated α-granules, pseudopodia, aggregation, and necrotic particles were observed in OxLDL alone and FH-HDL + OxLDL-treated platelets. In contrast, administration of control HDL prevented the features of OxLDL-induced platelet structural damage. Furthermore, activation of P-Selectin in response to an agonist ADP (50µM) was significantly inhibited by the HDL isolated from healthy subjects (ADP vs ADP + healthy HDL; 55.0±2.8 vs 20.0±2.8). Interestingly, the HDL isolated from FH subjects had no effect on ADP-induced P-Selectin activation (Figure 4D). Collectively these novel findings suggest that the HDL in FH subjects is dysfunctional with markedly impaired antiapoptotic and antithrombotic activity, which likely causes excessive platelet stimulation and thrombotic events.

**Figure 4.**
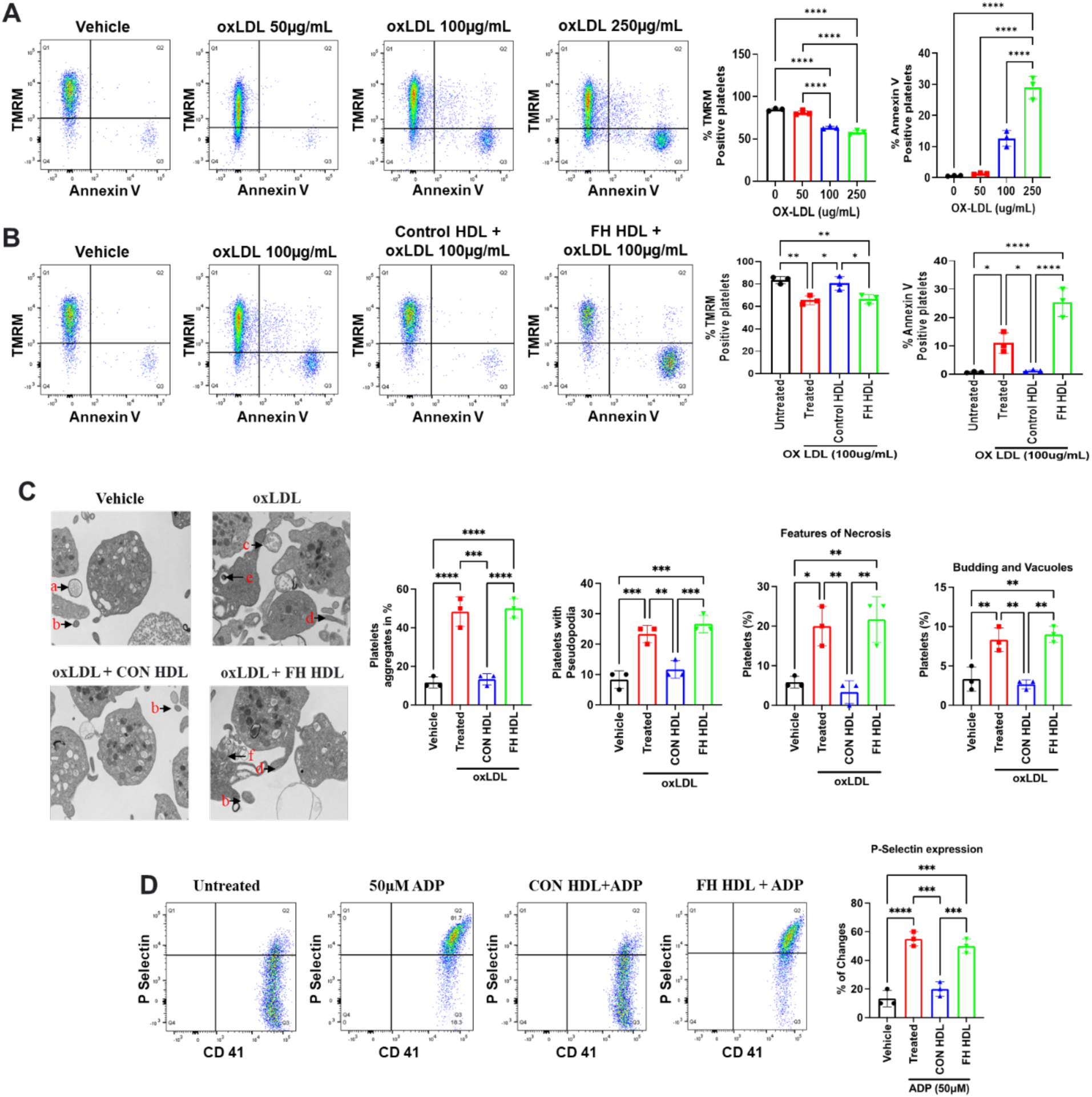
Loss of anti-apoptotic activity in HDL from FH patients. A, Healthy platelets exposed to varying concentrations of oxLDL (50, 100 and 250 µg/mL) for two hours. And the mitochondrial function and apoptosis were measured by flow cytometry using 40 nm TMRM and annexin V staining. **B,** 30 min preincubation with Control HDL (250 µg/mL) prevented the oxLDL (100 µg/mL)-induced apoptosis but FH-HDL (250 µg/mL) accelerated the apoptosis process in healthy platelets treated with oxLDL. **C,** Transmission electron microscopy also shows that FH- HDL increased the platelet aggregation, activity and necrosis in oxLDL-treated healthy platelets. (a) microsomes (b) platelets microparticle (c) budding (d) Pseudopodia (e) necrotic particle (f) content release from broken plasma membrane. **D,** Flow cytometry analysis revealed that 30min pretreatment with control HDL prevented the ADP (50µM)-induced platelets activation whereas FH-HDL did not alter the ADP mediated platelet activation. Data are expressed as mean fluorescence ± SEM. * - P ≤ 0.05; ** - P ≤ 0.01; *** - P ≤ 0.001.

### Evidence that FH-HDL accelerates platelet apoptotic signaling events

Since both activation and apoptosis occur in FH-HDL treated platelets, we next determined the OxLDL induced apoptotic signaling events in platelets treated with either control or FH-HDL (Figure 5). The importance of PKCδ as a crucial signaling mediator in the processes of platelet activation and aggregation cannot be overstated. Yet, the specific significance of PKCδ and its downstream effectors in the context of OxLDL-induced platelet apoptosis is still not fully understood. Treatment of OxLDL (100µg/mL) significantly increased PKCδ and their downstream proteins, activated Caspase 3 and Cytochrome C, in washed platelets. At the same time OxLDL treatment significantly downregulated the survival signaling proteins PI3K p85, pAKT (Thr 308) and BCL-xL in OxLDL treated platelets. Interestingly, FH-HDL (250µg/mL) treatment did not impact the proapoptotic or antiapoptotic signaling proteins in platelets treated with OxLDL. In contrast, HDL (250µg/mL) isolated from healthy subjects significantly decreased PKCδ, activated Caspase 3 and Cytochrome C levels and significantly increased PI3K p85, pAKT (Thr 308) and BCL-xL in OxLDL-treated platelets (Figure 5A). Our immunofluorescence staining also revealed that control HDL versus FH-HDL downregulated the proapoptotic proteins levels and increased antiapoptotic proteins levels in OxLDL treated platelets (Figure 5B).

**Figure 5.**
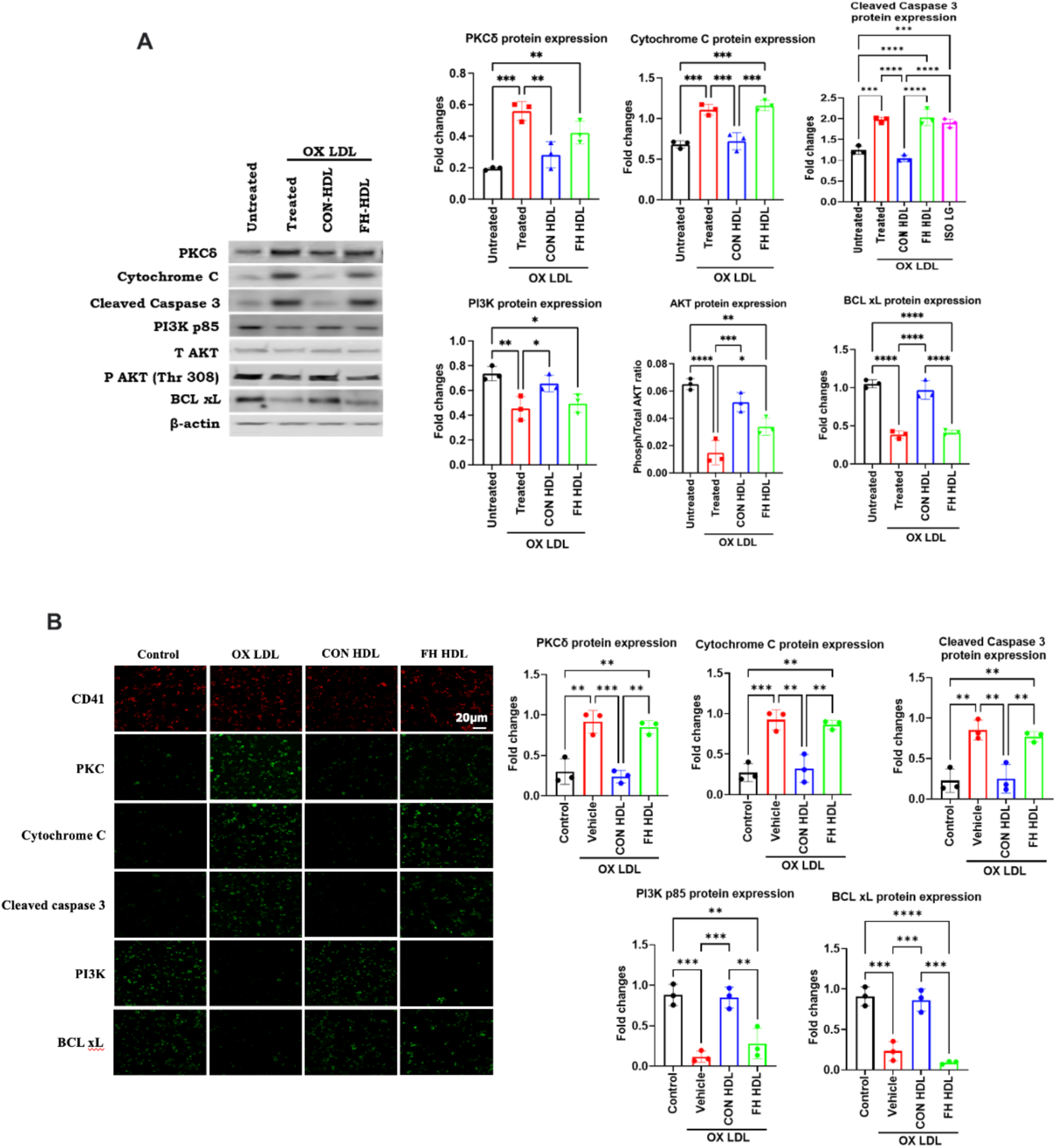
Differential impact of control and FH-HDL on OxLDL-induced platelet activation in healthy subjects. A, Platelets were pretreated with CON-HDL or FH-HDL (250 µg/mL) followed by exposure to OxLDL (100 µg/mL) and analyzed by western blot. **B,** Immunostaining was used to assess apoptosis and activation changes in platelets under the same treatment conditions. Data are expressed as mean ± SEM. * - P ≤ 0.05; ** - P ≤ 0.01; *** - P ≤ 0.001; **** - P ≤ 0.0001.

### Receptor dependent action of normal and dysfunctional HDL on platelet apoptosis

Lipoproteins act as ligands for the SR-BI and CD36 receptors, both of which are known to be present on the surface of platelets. Despite this, the capacity of these receptors on platelets to identify dysfunctional HDL remains unclear. To investigate this, we initially evaluated the interaction between control HDL and SR-BI by employing BLT1, a specific blocker of the SR-BI receptor (Figure 6A). For this study, platelets were pretreated with BLT1 and then incubated with oxLDL alone or with control HDL (Figure 6A). Due to BLT1 pretreatment, normal HDL lost its antiapoptotic activity. In this group, the proapoptotic proteins PKC δ, cytochrome C and activated caspase 3 levels were elevated and the antiapoptotic proteins PI3K and BCL-xL were decreased compared to the untreated group (normal HDL + OxLDL) (Figure 6A). There were no significant effects of platelet pretreatment with BLT1 on the proapoptotic impact of FH-HDL (Figure 6B). Therefore, we examined the effects of SSO (Sulfo N Succinimidyl Oleate, 25µM), a potent CD36 receptor blocker to uncover the role of dysfunctional FH-HDL (Figure 6B). In this experiment, we pretreated the platelets with SSO for 15 minutes and then incubated the platelets with either control or FH-HDL. Interestingly, treatment of platelets with FH-HDL alone significantly elevated the levels of proapoptotic proteins and downregulated the levels of antiapoptotic proteins. Importantly, pretreatment with CD36 receptor blocker significantly prevented the FH-HDL-induced platelet apoptosis, as indicated by the diminished levels of proapoptotic markers compared to treatment with FH-HDL alone. We also determined whether HDL/SRB1 downstream signaling activates the PI3K/BCL-xL pathway (Figure 6C), by pretreating the platelets with a PI3K inhibitor. Platelets treated with normal HDL + OxLDL exhibited decreased anti-apoptotic and increased proapoptotic protein effects with PI3K inhibition. Collectively, these data demonstrate that normal HDL binds with SRB1 receptor and activates PI3K, thereby inhibiting OxLDL-induced platelet apoptosis; whereas FH-HDL is compromised in mediating SR-BI activation of the PI3K/BCL-xL pathway. Furthermore, the dysfunctional FH-HDL triggers platelet apoptosis through CD36 receptor dependent downstream pathway.

**Figure 6.**
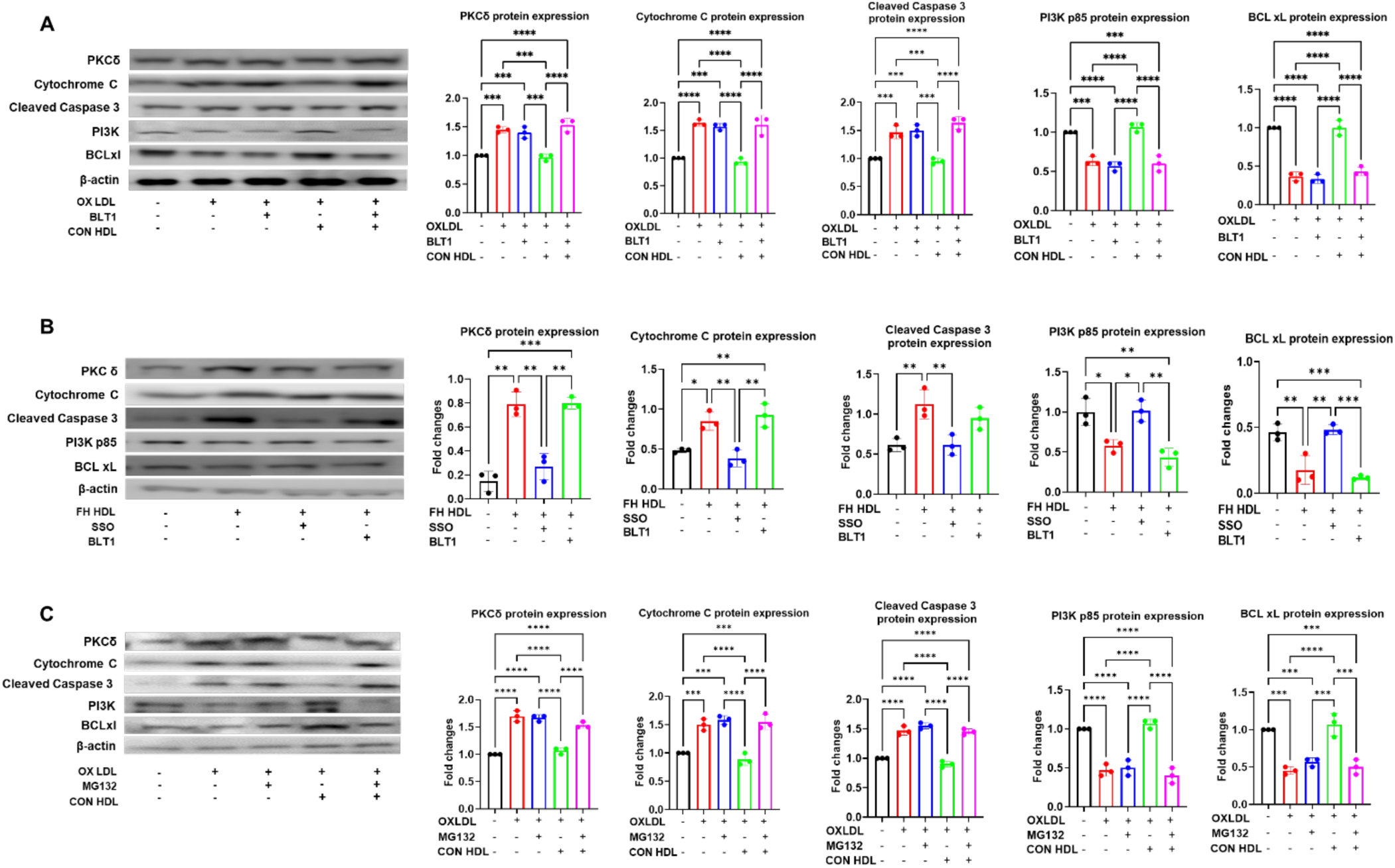
SR-B1 and CD36 receptor-mediated effects on HDL-induced apoptosis in platelets. A, SRB1 receptor-dependent anti-apoptotic effects of HDL. Western blot analysis showed that BLT1 (10nM), an SRB1 blocker prevented the protective role of HDL. In contrast, SR-B1 inhibition did not affect the oxLDL-induced apoptosis. **B,** FH-HDL induces platelet apoptosis through the CD36 receptor. CD36 inhibition by Sulfo-N-succinimidyl oleate (SSO, 25µM) prevented the FH-HDL-mediated apoptosis in platelets isolated from healthy subjects. **C,** Washed platelets were treated with MG 132 (10 µM) to inhibit the PI3K pathway and then cells were incubated with oxLDL or CON-HDL or FH-HDL. After the two hours treatment apoptotic proteins were measured. Data was expressed as mean ± SEM. * - P ≤ 0.05; ** - P ≤ 0.01; *** - P ≤ 0.001.

### Dicarbonyl scavengers reduce thrombosis formation in *Ldlr*^-/-^ knockout mice

FH subjects have higher levels of lipoprotein dicarbonyl adducts, which likely impacts the HDL anti-apoptotic capacity. In this *in vivo* study, we determined the effects of the dicarbonyl scavenger, 2-HOBA on thrombosis formation, occlusion time, and blood flow in *Ldlr***^-/-^** mice, which serve as a model for FH. Male *Ldlr*^-/-^ mice, aged six weeks, were subjected to a western diet for a duration of 16 weeks. During this period, they received continuous treatment with either plain drinking water or water infused with 2-HOBA at a concentration of 2g/L. Treatment with 2-HOBA significantly increased carotid artery occlusion time in *Ldlr^-/-^* mice (Figure 7A). Furthermore, the aldehyde scavenger prolonged the bleeding time in *Ldlr^-/-^*mice (Figure 7B). These findings indicate that 2-HOBA has antithrombotic effects in *Ldlr^-/-^* mice which is aligning with our earlier studies that showed 2-HOBA improves the atheroprotective functions of HDL in *Ldlr^-/-^* mice.

**Figure 7.**
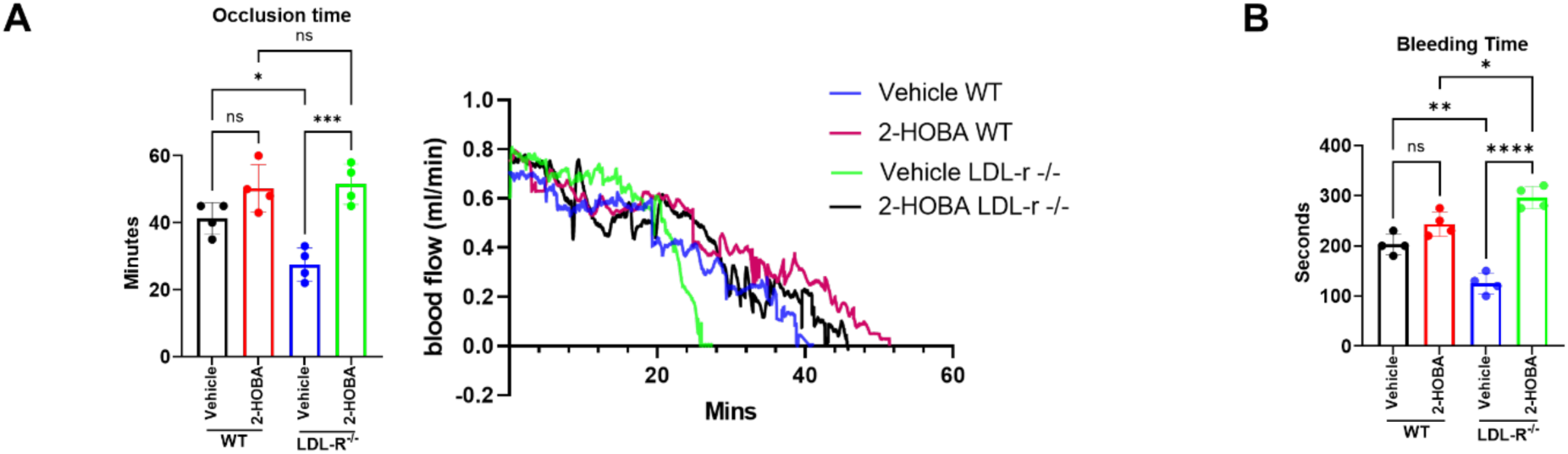
2-HOBA increased the occlusion time in *Ldlr^−/−^* mice. Eight-week-old male *Ldlr*^−/−^ mice underwent a two-week pretreatment with 1 g/L of 2-HOBA or a control vehicle (water). Following this initial phase, the treatments continued for an additional 16 weeks while the mice were maintained on a Western-style diet. **A,** After the treatment, the mice underwent carotid artery surgery, and blood flow was assessed using a Doppler flow probe. **B,** Additionally, the tail bleeding time was recorded in the *Ldlr*^−/−^ mice after the treatment period. N=10. Data was expressed as mean ± SEM. * - P ≤ 0.05; ** - P ≤ 0.01; *** - P ≤ 0.001.

## Discussion

Our previous studies suggest normal HDL can reduce agonist-stimulated platelet activation and aggregation^24^, while oxidized HDL enhances platelet aggregation, yet the mechanisms are not clear. In this study, we demonstrated that individuals with FH consistently show increased dysfunctional HDL-mediated thrombosis, likely due to enhanced platelet function and apoptosis. Mounting evidence supports the concept that dysfunctional HDL, for example, oxidized HDL, loses its beneficial properties and contributes to the development of cardiovascular disease. We have previously reported that HDL from patients with FH is enriched with lipid dicarbonyls, including MDA and IsoLG, and is strikingly dysfunctional^25,26^. Dysfunctional HDL has been associated with impairments in the regulation of platelet function. Normally, HDL helps maintain a balance in platelet activation to prevent excessive clot formation^27^. Our research reveals that the platelets in FH subjects are hyperactive. The FH- versus control-platelet structural alterations indicate platelets from FH subjects are more active and aggregated. The activated platelets generate microparticles of 0.1 to 1 μm in diameter that can transfer RNA and cell surface receptors to monocytes and endothelial cells, promoting platelet activation^28^. Increased platelet activation exacerbates this risk by accelerating thrombotic events on top of any underlying vessel damage caused by atherosclerosis. Dysfunctional HDL may fail to regulate platelet activity properly, leading to increased platelet hyperactivity. Indeed, we show that the dysfunctional HDL in FH subjects promotes platelet aggregation at a faster rate after addition of ADP, suggesting that FH-HDL contributes to the platelet hyperactivity in FH subjects.

OxLDL induced platelet activation triggers a cascade of events within the cell^29^. The process begins with the depolarization of the mitochondrial membrane, leading to an influx of calcium into the cytoplasm. The surge in this influx causes the mitochondrial permeability transition pore (MPTP) to open, which ultimately leads to a decrease in the potential of the inner mitochondrial membrane. These early mitochondrial changes precede subsequent cellular responses, including the translocation of phosphatidylserine (PS) to the outer leaflet of the platelet membrane. This externalization is a marker of platelet activation and also promotes platelet adhesion and apoptosis^30^. We currently show that unlike control HDL, HDL from FH subjects is markedly impaired in preventing platelet mitochondrial membrane depolarization in response to oxLDL. Morphologically, platelet apoptosis manifests as platelet shrinkage, externalization of PS on the outer membrane, and fragmentation into microparticles. The regulation of platelet survival is largely governed by Bcl-xL, an essential inhibitor of BAK and BAX, two pro-apoptotic proteins that target mitochondria for damage. Additionally, PKCδ facilitates cell death by either inhibiting the protective Bcl-2 family proteins or enhancing the expression of proapoptotic BH3 domain-containing proteins, thereby promoting the creation of Bax/Bak channels, cytochrome c release, and caspase activation^31^.

In our investigation, we observed upregulated pro-apoptotic proteins level and downregulated anti-apoptotic proteins level in platelets from FH compared to healthy individuals. When platelets encounter stressors like oxLDL, the usual survival signals are overwhelmed. This triggers the activation of PKCδ, initiating formation of Bax/Bak channel and the cytochrome c release through pores in the mitochondrial membrane. In the cytosol, cytochrome c then initiates the apoptotic caspase cascade, causing the cleavage of numerous cellular constituents and ultimately disrupting vital cellular processes. In this regard, FH subjects have markedly elevated levels of oxLDL. Furthermore, we show that FH versus control HDL is not only ineffective at preventing oxLDL induced platelet apoptosis, but FH-HDL alone is a trigger of platelet apoptosis (Figure 6B). This intricate interplay underscores the critical role of mitochondrial-mediated apoptosis in platelet biology and its implications in cardiovascular diseases associated with dyslipidemia and enhanced oxidative burden^31^.

Numerous mechanisms have been suggested to explain the anti-apoptotic properties of HDL in cells, which vary based on the specific triggers of apoptosis. For instance, OxLDL induces a gradual yet prolonged rise in intracellular calcium levels within endothelial cells, ultimately resulting in cell death. However, this detrimental effect can be counteracted by HDL, which works by inhibiting the increase in calcium levels^32^. HDL plays a crucial role in preventing apoptosis in endothelial cells induced by TNF-α. This protective effect is linked to a reduced activation of caspase 3, a key player in the primary apoptotic pathways. ^33^. HDL typically acts as a protective agent for LDL by absorbing lipid peroxides and their reactive by-products, preventing harmful aldehyde adduction. However, when HDL undergoes modification, it loses many of its essential functions, such as cholesterol efflux, anti-inflammatory properties, and the activation of PON1.^25,26^. Platelets that experience a significant loss of mitochondrial membrane potential fail to react to agonist stimulation. In reality, the process of apoptosis leads to a heightened turnover of platelets, resulting in the production of larger, younger platelets that exhibit greater activity, ultimately contributing to an increased risk of thrombosis. Several antithrombotic mechanisms that protect against platelet hyperactivity have been suggested in the literature for HDL^31^. Our BLT1 inhibitor study clearly indicates that the HDL binds to SB-B1 receptor leading to PI3K and Akt activation (Figure 8). Activation of Akt also phosphorylates the Bcl-2-associated death promoter (Bad), thereby inhibiting its interaction with Bcl-xL. This allows Bcl-xL to suppress mitochondria-dependent apoptosis process. At the same time, dysfunctional HDL binds with CD36 receptor promoting the activation of PKCδ mediated intrinsic pathway. Upon activation, PKCδ may translocate to the mitochondria and interact with Bcl-2 family proteins like BAK and Bax, which may lead to the permeabilization of the mitochondrial outer membrane. This triggers the cytochrome c release along with other pro-apoptotic proteins from the mitochondria, leading to caspase activation and apoptosis^30^.

**Figure 8.**
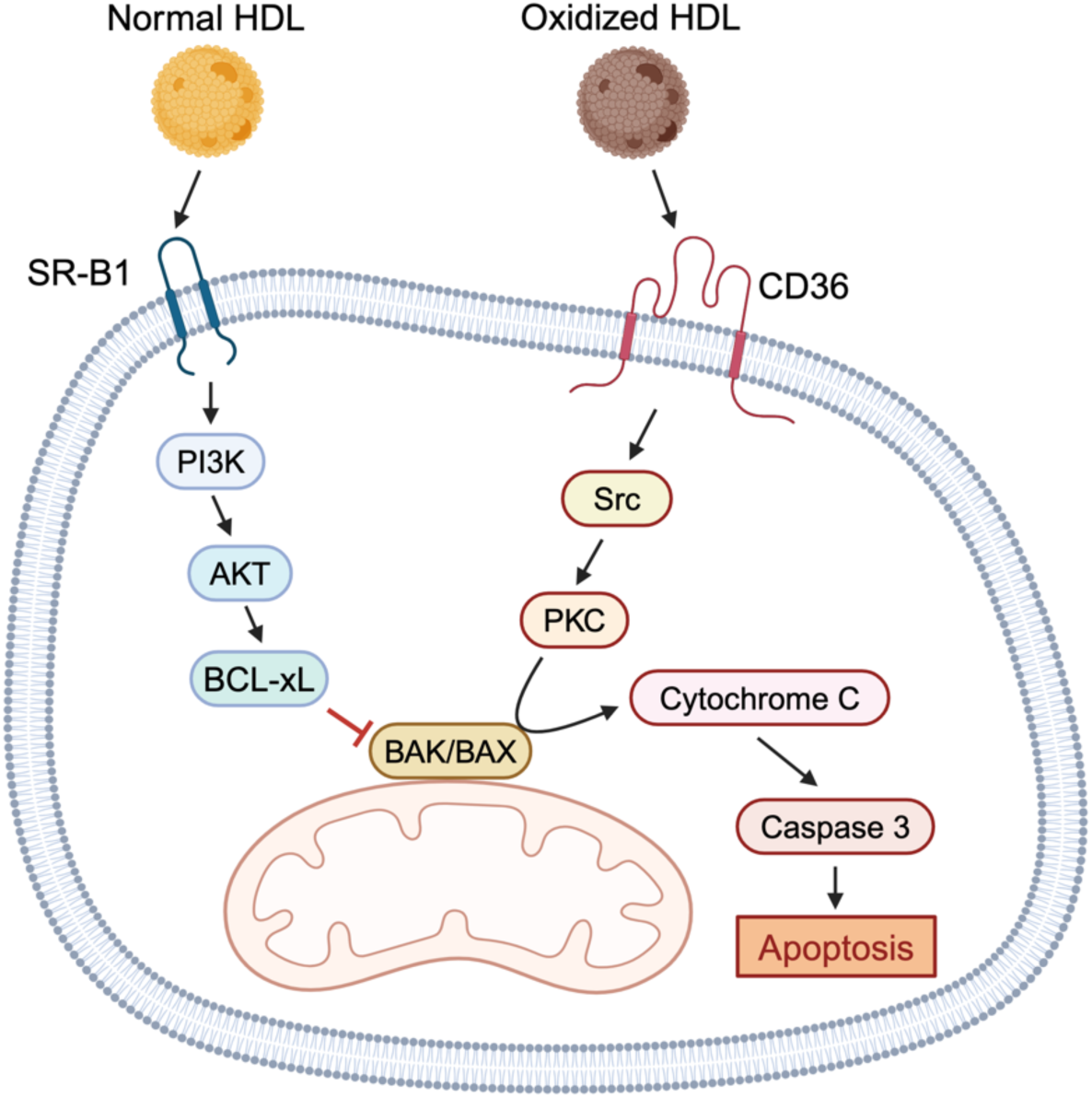
Mitochondrial depolarization as an early apoptosis indicator in platelet activation and thrombotic risk. Mitochondrial depolarization serves as a key indicator of early apoptosis, closely linked to the release of pro-apoptotic elements such as cytochrome C, which in turn activate caspases and set off a series of apoptotic processes. Our findings indicate that normal HDL interacts with the SR-B1 receptor, enhancing its anti-apoptotic functions. Conversely, the binding of oxidized LDL to the CD36 receptor leads to platelet activation and aggregation, which can significantly increase the risk of thrombotic events. The figure was prepared using BioRender.com.

Novel aldehyde scavengers, which counteract the adverse consequences of reactive oxidative stress (ROS) while preserving the essential signaling of ROS, can reduce the dicarbonyl modification of the HDL and prevent formation of dysfunctional HDL^25,34^. Research indicates that 2-HOBA, a dicarbonyl scavenger, has the potential to diminish the modification and dysfunction of HDL caused by MDA and IsoLG. We previously reported that the 2-HOBA plays a crucial role in significantly slowing down the progression of atherosclerosis in the hypercholesterolemic *Ldlr*^−/−^ mouse model of FH. Notably, the treatment with 2-HOBA has been shown to enhance the stability of atherosclerotic plaques, as demonstrated by a reduction in necrosis and an increase in both fibrous cap thickness and collagen levels^25^. Consistent with our present findings, dicarbonyl scavenging decreases MDA modification in HDL *in vivo* and improves its antithrombotic activity in *Ldlr^−/−^* mouse. 2-HOBA treatment significantly increased the occlusion and bleeding time in *Ldlr^−/−^*mice. Employing 2-HOBA for dicarbonyl scavenging offers a promising therapeutic strategy to slow the progression of atherosclerosis and reduce the risk of clinical events linked to the formation of unstable atherosclerotic plaques.

## Conclusion

HDL’s health-promoting impact on the cardiovascular system have been attributed to its capacity to improve reverse cholesterol transport along with its antioxidant, anti-inflammatory, and antithrombotic properties. HDL’s antithrombotic effect may involve direct modulation of platelet function. However, studies have yielded conflicting results regarding HDL’s impact on activation and aggregation of platelets. While most research supports HDL playing a crucial role in reducing platelet activation, some studies suggest either increased activation or no significant effect. Interestingly, recent studies demonstrated that apoptosis also occurs in anucleate platelets, and apoptotic platelets accelerate in vivo arterial thrombosis formation. Mitochondrial depolarization is a hallmark of early apoptosis and is associated with the efflux of pro-apoptotic proteins like cytochrome c, which triggers caspases and initiates a cascade of events that result in apoptosis.

Our data suggest that normal HDL binds with the SR-B1 receptor, promoting its antiapoptotic activity, while OxLDL/CD36 binding triggers platelet activation and aggregation, thereby facilitating thrombotic events.

## Nonstandard Abbreviations and Acronyms

FH: familial hypercholesterolemia
oxLDL: oxidized LDL
Ldlr: LDL receptor
2-HOBA: 2-hydroxybenzylamine
BLT1: Block lipid transport-1
SSO: sulfo-N-succinimidyl oleate
Apo A1: Apolipoprotein A1
PS: phosphatidylserine
PRP: Platelet-rich plasma

## Data Availability

All data supporting the findings of this study are available within the article and its supplementary information files. Additional raw data, including RNA sequencing datasets and processed values, are available from the corresponding author upon reasonable request.

## Novelty and Significance

### What Is Known?

- HDL protects against cardiovascular disease through cholesterol removal and anti-inflammatory actions.
- Dysfunctional HDL impairs cholesterol efflux and promotes inflammation, increasing cardiovascular risk.
- Traditional therapies aimed at increasing HDL quantity have failed to reduce cardiovascular events, shifting focus to improving HDL quality and function.

### What New Information Does This Article Contribute?

- Our study reveals specific peroxidation products in HDL from FH patients that promote thrombosis.
- We identify abnormal interactions between dysfunctional HDL and the SR-B1 and CD36 receptors, contributing to cardiovascular complications in FH.
- We demonstrate that 2-HOBA mitigates the pro-thrombotic effects of dysfunctional HDL, offering a new therapeutic strategy for FH and the treatment of atherosclerosis.

### Summary

Our research provides significant advancements in understanding the role of dysfunctional HDL in FH. We have identified key peroxidation products that contribute to the pro-thrombotic nature of HDL in FH patients, delineating a clear pathogenic mechanism through which HDL exacerbates platelet activation and thrombosis. Importantly, we show that the intervention with 2-HOBA can reverse these harmful effects, offering a promising therapeutic approach. This study not only underscores the complexity of HDL functionality in cardiovascular disease but also opens new avenues for targeted treatments that could benefit individuals with FH, who are at high risk for thrombotic events.

